# Impact of Insurance Status on Late-Stage Disease Presentation and Disease-Specific Survival among US Patients With Gastric Cancer

**DOI:** 10.1101/2023.12.26.23300531

**Authors:** Ted O. Akhiwu, Jincong Q. Freeman, Adam W. Scott, Victoria Umutoni, Philip O. Kanemo

## Abstract

**Purpose:** The impact of insurance status on cause-specific survival and late-stage disease presentation among US patients with gastric cancer (GC) has been less well-defined.

**Materials and Methods:** A retrospective study analyzed the 2007-2016 Surveillance Epidemiology and End Results. GC events were defined as GC-specific deaths; patients without the event were censored at the time of death from other causes or last known follow-up. Late-stage disease was stage III-IV. Insurance status was categorized as “uninsured/Medicaid/private.” Five-year survival rates were compared using log-rank tests. Cox regression was used to assess the association between insurance status and GC-specific survival. Logistic regression was used to examine the relationship of insurance status and late-stage disease presentation.

**Results:** Of 5,529 patients, 78.1% were aged ≥50 years; 54.2% were White, 19.4% Hispanic, and 14.0% Black; 73.4% had private insurance, 19.5% Medicaid, and 7.1% uninsured. The 5-year survival was higher for the privately insured (33.9%) than those on Medicaid (24.8%) or uninsured (19.2%) (p<0.001). Patients with Medicaid (adjusted hazard ratio [aHR] 1.22, 95%CI: 1.11-1.33) or uninsured (aHR 1.43, 95%CI: 1.25-1.63) had worse survival than those privately insured. The odds of late-stage disease presentation were higher in the uninsured (adjusted odds ratio [aOR] 1.61, 95%CI: 1.25-2.08) or Medicaid (aOR 1.32, 95%CI: 1.12-1.55) group than those with private insurance. Hispanic patients had greater odds of late-stage disease presentation (aOR 1.35, 95%CI: 1.09-1.66) than Black patients.

**Conclusions:** Findings highlight the need for policy interventions addressing insurance coverage among GC patients and inform screening strategies for populations at risk of late-stage disease.

## Introduction

In the US, gastric cancer (GC) has an incidence of 1.5%, with over 26,000 patients newly diagnosed with GC every year and over 11,000 GC related deaths annually.^1,2,3^ In fact, GC is the sixth most common cancer worldwide, with GC accounting for up to 90% of all cases.^1,2^ Cigarette smoking, alcohol consumption, high salt diet, obesity, lack of physical activity, and *Helicobacter pylori* infection have all been associated as risk factors to the development of GC.^4^ Older age, male sex, advanced stage at diagnosis, and poor histologic grade have also found to contribute to poor survival outcomes in patients with GC.^5^ The highest incidence rates of GC have been reported in East Asian countries including China, Korea, and Japan. Additionally, within the US, Asian or Pacific Islander populations also experience the highest GC incidence among any minority ethnic group.^6^ In developing countries, GC has a high proportionate mortality and is only exceeded by lung, colon, and liver cancers.^7^ Overall, GC carries a poor prognosis with an overall 5-year survival rate of 32% in the US and 30% in Europe.^8,9^ However, some US racial and ethnic minority patients with GC have been shown to have poorer survival outcomes than Asian or Pacific Islander patients, despite having lower GC incidence rates.^10,11^

Over the past decades, rates of the uninsured US populations have ranged from 10%-20%, with a higher proportion of racial and ethnic minority populations being uninsured, notably Hispanic and Black populations.^13,14^ Insurance status serves as one reflection of a person’s socioeconomic status and has an important function in offsetting financial difficulties that might affect overall cancer diagnosis and/or survival, including access to rehabilitation and follow-up visits. Studies of the influence of insurance status on multiple cancers including those of the salivary gland, breast, brain, and bone have shown worse health outcomes and 5-year survival rates for the uninsured than for the insured populations.^15–18^ For example, a study found that insured patients had an 8.5% higher 5-year cause-specific survival than uninsured patients.^19^ Another study conducted by Findakly et al. also reported worse survival outcomes in GC patients who were on Medicaid and those without insurance compared to those privately insured.^20^

The stage of GC at diagnosis constitutes another risk factor affecting survival outcomes in cancer patients. It is not surprising that systemic barriers to health insurance and access to healthcare can delay preventive screening, diagnosis, and treatment for patients with GC. One analysis found that among 249 primarily immigrant patients diagnosed or treated at an urban academic safety net hospital, patients with later stage of disease presentation (stages III-IV) had a significantly lower survival advantage than those with earlier stage (stage I-II).^21^ Additionally, significant associations with late-stage presentation were found among patients that identified as having a Hispanic ethnicity, Medicaid insurance (as opposed to private insurance), and neighborhood-level poverty. Since GC survival rates are higher at localized stage of the disease, early detection of many cancers could confer a improved survival advantage as opposed to delayed detection, and this could be influenced by the insurance coverage a patient has. Previous research on the association of insurance status and GC outcomes have shown worse survival outcomes while others have documented no difference in the rate of survival.^19–21^ However, the sample sizes of these studies may not have been adequately powered to confidently assert these conclusions. In addition, none of these studies focused on the worst histologic type of GC, gastric adenocarcinoma.^20,21,23^ Overall, the association between insurance status and GC-specific survival has not been well characterized particularly among US patients.

The primary goal of this study was to examine the association between insurance status and cause-specific survival among patients aged 18-64 years who were diagnosed with GC. The secondary goal was to assess the impact of insurance status on the stage of disease presentation by comparing the risk of advanced stage disease between insured and uninsured patients.

## Materials and Methods

### Study design, population, and data source

This study was a retrospective cohort study. Data from the November 2018 submission of the Surveillance Epidemiology and End Results (SEER) 18 Registry (1975-2016) was used for this study as more recently updated submissions (November 2019 and 2020) had the insurance recode variable removed to avoid the risk of re-identification.^24^ The SEER database collects incidence, mortality, and survival data from population-based cancer registries across the nation, encompassing 28% of the US general population.^24^ This database was chosen because it is publicly available using the SEER*Stat Software and contains sufficient statistics on sociodemographic and tumor characteristics and survival outcomes. A total of 10,664 patients with GC were initially identified, and the final sample size after applying the inclusion and exclusion criteria was 5,529.

Patients with biopsy-proven GC between the ages of 18-64 years were included in this analysis. Information on patients with pathologically confirmed GC was extracted from the SEER database by anatomic sites using the International Classification of Diseases for Oncology (ICD-O-3) code for the stomach: C16.0–16.9, and by histologic codes for GC (8140). Data on insurance status became available in 2007, so this study utilized data collected from patients diagnosed with GC between 2007 and 2016. Exclusion criteria included patients ages 65+ in whom insurance status was difficult to ascertain due to Medicare eligibility; with unknown insurance status; with unknown disease staging; and/or with missing follow-up time. This study was exempt from our Institutional Review Board because it was conducted on publicly available de-identified data.

### Study variables

The main independent variable was insurance status categorized into three groups: Uninsured, Medicaid, and insured (commercial or private insurance). Covariates assessed included age at the time of diagnosis, sex, race/ethnicity, marital status, American Joint Committee on Cancer (AJCC-TNM) tumor stage, year of diagnosis, and survival months. Race/ethnicity was categorized as Non-Hispanic Black, Non-Hispanic White, Hispanic, and Other. Marital status is coded as: single or never married, not married (divorced, widowed, separated, unmarried domestic partner), and unknown. The AJCC stage consisted of stage I, II, III, and IV. The primary outcome in this study was the cause-specific survival of patients with GC. The secondary outcome was the odds of late-stage presentation (defined as stage III-IV) in relation to participants’ insurance status.

### Statistical analysis

Baseline characteristics were summarized and presented as frequencies (percentages [%]) for categorical variables and means (standard deviations [SD]) for continuous variables. The distributions of characteristics across insurance groups were compared using ANOVA for continuous data and Pearson’s chi-squared tests for categorical data. Simple logistic regression was used to model the odds of late-stage disease presentation on insurance status, followed by multiple logistic regression adjusting for potential confounders. Crude odds ratios, adjusted odds ratios (aOR), and 95% confidence intervals (95% CI) were calculated.

Events were defined as GC-specific cancer-related mortality. Patients who did not experience the event were censored at the time of their death from other causes or their last known follow-up visit or contact. Kaplan-Meier curves of GC-specific survival across for insurance groups and median survival time were compared using log-rank tests. Cox proportional hazards regression models were fit to estimate adjusted hazard ratios (aHR) and 95% CIs for cause-specific survival stratified by insurance status. Covariates included age, sex, race/ethnicity, marital status, and AJCC stage. An interaction term of race/ethnicity and insurance status was applied to assess the interaction between the two variables; however, no statistically significant interaction between race/ethnicity and insurance status (*P*=0.896) was observed. *P*-values were reported as two-sided at the 0.05 level of significance. All statistical analyses were performed using SAS 9.4 (SAS Institute, Cary, NC).

## Results

Of 5,529 patients, 4,057 (73.4%) had private insurance, 1,077 (19.5%) were on Medicaid, and 395 (7.1%) were uninsured. The overall mean age at diagnosis was 54.6 (SD=8.1) years. Higher proportions of the patients were male (73.7%) and non-Hispanic White (54.2%). There were statistically significant differences in the distributions of age at diagnosis (*P*<0.001), sex (*P*=0.020), race/ethnicity (*P*<0.001), marital status (*P*<0.001), and AJCC stage *P*<0.001) by insurance status (**Table 1**).

**Table 1.** Demographic and clinical characteristics of patients with gastric cancer, overall and by insurance status.

|  | **Overall**<br>**(N=5,529)** | **Insured**<br>**(n=4,057 [73.4%])** | **Medicaid**<br>**(n=1,077 [19.5%])** | **Uninsured**<br>**(n=395 [7.1%])** | **P value \*** |
| --- | --- | --- | --- | --- | --- |
| **Age at diagnosis, mean (SD)** | 54.6 (8.1) | 55.1 (7.9) | 53.2 (8.5) | 53.1 (8.8) | <0.001 |
| **Age at diagnosis, median (IQR)** | 56 (51-61) | 57.0 (51.0-61.0) | 55.0 (49.0-60.0) | 55.0 (49.0-60.0) | <0.001 |
| **Survival months, median (IQR)** | 14.0 (6.0-28.0) | 15.0 (7.0-31.0) | 11.0 (4.0-22.0) | 6.0 (2.0-17.0) | <0.001 |
| **Age group (year), n (%)** |  |  |  |  |  |
| <50 | 1,211 (21.9) | 802 (19.8) | 302 (28.0) | 107 (27.1) | <0.001 |
| 50+ | 4,318 (78.1) | 3,255 (80.2) | 775 (72.0) | 288 (72.9) |  |
| **Sex, n (%)** |  |  |  |  |  |
| Male | 4,075 (73.7) | 3,027 (74.6) | 758 (70.4) | 290 (73.4) | 0.020 |
| Female | 1,454 (26.3) | 1,030 (25.4) | 319 (29.6) | 105 (26.6) |  |
| **Race/Ethnicity, n (%)** |  |  |  |  |  |
| White | 2,999 (54.2) | 2,455 (60.5) | 391 (36.3) | 153 (38.7) | <0.001 |
| Black | 772 (14.0) | 520 (12.8) | 186 (17.3) | 66 (16.7) |  |
| Hispanic | 1,071 (19.4) | 614 (15.1) | 330 (30.6) | 127 (32.2) |  |
| Other | 687 (12.4) | 468 (11.5) | 170 (15.8) | 49 (12.4) |  |
| **Marital status, n (%)** |  |  |  |  |  |
| Married | 3,303 (59.7) | 2,693 (66.4) | 435 (40.4) | 175 (44.3) | <0.001 |
| Not married | 1,987 (35.9) | 1,206 (29.7) | 583 (54.1) | 198 (50.1) |  |
| Unknown | 239 (4.3) | 158 (3.9) | 59 (5.5) | 22 (5.6) |  |
| **Tumor stage, n (%)** |  |  |  |  |  |
| I | 822 (14.9) | 637 (15.7) | 141 (13.1) | 44 (11.1) | <0.001 |
| II | 759 (13.7) | 595 (14.7) | 125 (11.6) | 39 (9.9) |  |
| III | 1,316 (23.8) | 986 (24.3) | 253 (23.5) | 77 (19.5) |  |
| IV | 2,632 (47.6) | 1,839 (45.3) | 558 (51.8) | 235 (59.5) |  |

Abbreviations: SD = standard deviation; IQR = interquartile range.
\* P values were computed using ANOVA, Kruskal-Wallis, or Chi-square tests as appropriate.

In **Table 2**, after covariate adjustment, the odds of late-stage presentation were significantly higher among patients without insurance (aOR, 1.61; 95% CI, 1.25-2.08) or on Medicaid (aOR, 1.32; 95% CI, 1.12-1.55) compared to those privately insured. Hispanic patients had a significantly greater likelihood of late-stage disease presentation than non-Hispanic Black patients (aOR, 1.50; 95% CI, 1.22-1.85), and this disparity persisted in multivariable analysis (aOR, 1.35; 95% CI, 1.09-1.66). Persons who were 50+ years had a significantly lower odds of presenting late-stage disease at diagnosis compared with those aged 50 years (aOR, 0.66; 95% CI, 0.57-0.77).

**Table 2.** Logistic regression modeling for late-stage disease (stages III-IV) in patients with gastric cancer.

|  | **OR (95% CI)** | **P value** | **aOR<sup>a</sup> (95% CI)** | **P value \*** |
| --- | --- | --- | --- | --- |
| **Insurance status** |  |  |  |  |
| Insured | 1.0 (reference) |  | 1.0 (reference) |  |
| Medicaid | 1.33 (1.14-1.55) | <0.001 | 1.32 (1.12-1.55) | <0.001 |
| Uninsured | 1.64 (1.28-2.11) | <0.001 | 1.61 (1.25-2.08) | <0.001 |
| **Sex** |  |  |  |  |
| Female | 1.0 (reference) |  | 1.0 (reference) |  |
| Male | 1.09 (0.95-1.24) | 0.219 | 1.12 (0.98-1.28) | 0.093 |
| **Race/Ethnicity** |  |  |  |  |
| Black | 1.0 (reference) |  | 1.0 (reference) |  |
| White | 1.11 (0.94-1.32) | 0.229 | 1.14 (0.95-1.35) | 0.156 |
| Hispanic | 1.50 (1.22-1.85) | <0.001 | 1.35 (1.09-1.66) | 0.006 |
| Other | 1.15 (0.92-1.44) | 0.227 | 1.09 (0.87-1.37) | 0.448 |
| **Age group (year)** |  |  |  |  |
| <50 | 1.0 (reference) |  | 1.0 (reference) |  |
| 50+ | 0.64 (0.55-0.75) | <0.0001 | 0.66 (0.57-0.77) | <0.001 |
| **Marital status** |  |  |  |  |
| Married | 1.0 (reference) |  | 1.0 (reference) |  |
| Not married | 0.96 (0.85-1.08) | 0.481 | 0.90 (0.79-1.02) | 0.108 |
| Unknown | 0.77 (0.59-1.02) | 0.073 | 0.74 (0.56-0.99) | 0.039 |

Abbreviations: OR = odds ratio; aOR = adjusted odds ratio; CI = confidence interval.
\* adjusted for all variables in the table.

The Kaplan-Meier curve for cause-specific survival as a function of insurance status is illustrated in **Figure 1**. Patients with private insurance had a longer median survival time (22.0; 95% CI, 21.0-23.0) compared to those on Medicaid (14.0; 95% CI, 13.0-16.0) and those uninsured (10.0; 95% CI, 8.0-12.0). The 5-year cause-specific rate of survival was also significantly higher for privately insured patients (33.9%) than for those on Medicaid (24.8%) and those uninsured patients (19.2%) (*P*<0.001).

**Figure 1.**
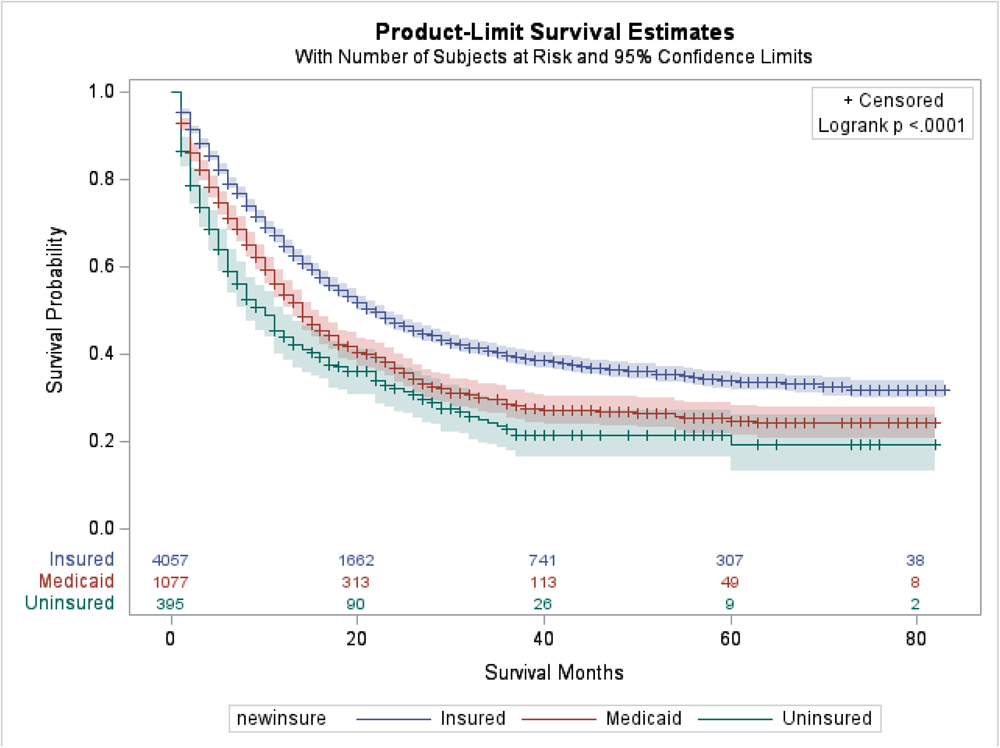
Kaplan-Meier curve displaying cause-specific disease survival as a function of insurance status among patients with gastric cancer.

The adjusted Cox regression results (**Table 3**) showed that having Medicaid (aHR, 1.22; 95% CI, 1.11-1.33), no insurance (aHR, 1.43; 95% CI, 1.25-1.63), male sex (aHR, 1.11; 95% CI, 1.02-1.20), and being unmarried (aHR, 1.18; 95% CI, 1.09-1.27) were significantly associated worse cause-specific survival. The stage at disease presentation also exponentially increased the risk of poor survival. As the stage of disease presentation progressed from stage I to stage IV, GC-specific mortality increased significantly (stage II: aHR, 2.38; 95% CI, 1.93-2.94); stage III: aHR, 4.28; 95% CI, 3.55-5.17; stage IV: aHR, 12.57; 95% CI, 10.50-15.04). No statistically significant difference in mortality by race/ethnicity was observed (**Table 3**).

**Table 3.** Cox proportional hazards model for cause-specific survival among patients with gastric cancer.

|  | **HR (95% CI)** | **P value** | **aHR<sup>a</sup> (95% CI)** | **P value \*** |
| --- | --- | --- | --- | --- |
| **Insurance status** |  |  |  |  |
| Insured | 1.0 (reference) |  | 1.0 (reference) |  |
| Medicaid | 1.36 (1.25-1.48) | <0.001 | 1.22 (1.11-1.33) | <0.001 |
| Uninsured | 1.72 (1.51-1.96) | <0.001 | 1.43 (1.25-1.63) | <0.001 |
| **Age group (year)** |  |  |  |  |
| <50 | 1.0 (reference) |  | 1.0 (reference) |  |
| 50+ | 0.84 (0.78-0.92) | <0.001 | 1.003 (0.92-1.09) | 0.946 |
| **Sex** |  |  |  |  |
| Female | 1.0 (reference) |  | 1.0 (reference) |  |
| Male | 1.07 (0.98-1.15) | 0.127 | 1.11 (1.02-1.20) | 0.015 |
| **Race/Ethnicity** |  |  |  |  |
| Black | 1.14 (1.03-1.26) | 0.015 | 1.06 (0.95-1.17) | 0.309 |
| White | 1.0 (reference) |  | 1.0 (reference) |  |
| Hispanic | 1.23 (1.12-1.34) | <0.001 | 1.07 (0.98-1.18) | 0.146 |
| Other | 1.03 (0.92-1.15) | 0.636 | 1.01 (0.90-1.13) | 0.869 |
| **Marital status** |  |  |  |  |
| Married | 1.0 (reference) |  | 1.0 (reference) |  |
| Not married | 1.21 (1.13-1.31) | <0.001 | 1.18 (1.09-1.27) | <0.001 |
| Unknown | 0.91 (0.75-1.09) | 0.284 | 0.88 (0.73-1.06) | 0.178 |
| **Stage** |  |  |  |  |
| I | 1.0 (reference) |  | 1.0 (reference) |  |
| II | 2.43 (1.96-3.00) | <0.001 | 2.38 (1.93-2.94) | <0.001 |
| III | 4.34 (3.59-5.23) | <0.001 | 4.28 (3.55-5.17) | <0.001 |
| IV | 12.82 (10.72-15.34) | <0.001 | 12.57 (10.50-15.04) | <0.001 |

Abbreviations: HR = hazard ratio; aHR = adjusted hazard ratio; CI = confidence interval.
\* Adjusted for demographic characteristics and tumor stage

## Discussion

Our population-based study demonstrated that insurance status had a significant association with the cause-specific survival outcomes among patients with GC and late-stage disease presentation. Privately insured patients had a better GC-specific survival than those who were uninsured or on Medicaid, even after adjusting for tumor characteristics and sociodemographic factors. Similarly, patients without insurance or on Medicaid were more likely to present late-stage disease at diagnosis compared with those privately insured. Furthermore, race/ethnicity seemed not to modify the association between insurance status and GC-specific survival of patients in this study.

To our knowledge, the impact of insurance status on GC-specific survival has not been clearly clarified. Although some studies noted insurance status to be a predictive factor of poor survival outcomes in patients with GC,^15–18^ these studies were not limited to the GC subtype which is the more common and lethal subtype. We found that patients with private insurance had better survival outcomes compared to those who were uninsured or on Medicaid. Jin et al. observed similar results, when investigating the survival gap in GC between non-Hispanic Asian and White patients in the US, which found that private insurance and higher socioeconomic status significantly improved GC patients’ prognosis.^25^ However, their analysis included all histologic subtypes of GC including distal squamous and adenosquamous carcinomas, whereas our study focused solely on GC.

Findakly et al. conducted a large single-institution retrospective cohort study evaluating the impact of sociodemographic characteristics on GC outcomes among 111 patients with GC, demonstrating that lack of insurance or having Medicaid were associated with significantly worse survival outcomes.^20^ However, most of the patients in the study were uninsured, where fewer than 30% of them had any form of insurance. Another study by Jang et al., aimed to evaluate the association of medical insurance status of 333 Korean patients with GC and their survival after gastrectomy, reporting that the medical aid group had a similar survival rate compared to the Medicaid/Uninsured group.^26^ The median post-op duration of hospitalization was higher for the Medicaid group compared to the National Health insured group and the overall 5-year survival was significantly higher in the National Health Insurance registered group.^26^ In contrast, this study also analyzed differences in the distribution or level of factors that affect the prognosis of GC patients including the Eastern Cooperative Oncology Group scale score and comorbidities which are missing from the SEER database.

Health insurance has generally been known to facilitate greater access to health care, novel therapies, and increase the use of care services.^27^ For example, pre-operative chemotherapy among patients with resectable cT2 GC or higher has been shown to improve survival than surgery alone. However, despite this benefit of chemotherapy, significant disparities existed in the rate of utilization based on insurance status, with uninsured/Medicaid patients having lower odds of receiving pre-op chemotherapy.^28^ Additionally, Medicaid and dual-eligible patients with GC also have lower odds of having the recommended number of affected lymph nodes removed and examined compared with those privately insured, even after adjusting for known confounders.^29^ These could be potential explanations for the prolonged survival time noted in privately insured patients. Moreover, insurance status is an indirect indicator of socioeconomic status, and it is likely that individuals with excellent social support and financial capacity had better access to health care and services which might have accounted for their improved survival.^17^

The current study also showed that being uninsured or having Medicaid was associated with a higher likelihood of late-stage disease presentation, even after adjusting for age, race/ethnicity, sex, and marital status. These results are similar to those from an analysis performed by Morgan et al. that assessed the presentation and survival of GC patients at an urban safety-net facility in which having Medicaid was independently associated with late-stage disease presentation.^22^ Interestingly, in their study, lack of insurance was not significantly associated with late stage of disease presentation. One explanation could be the small sample size of the uninsured group, having led to inadequate power to detect a statistically significant difference. Contrarily, Robbins et al. assessed the relationship between insurance status and distant-stage disease and showed that insurance status was a strong predictor for distant disease presentation.^30^ However, this study used the National Cancer Database; many types of cancers other than GC were included in the analysis, with participants being restricted to adolescents and young adults only.

It is worthy to note that GC does not have a high incidence in the US to justify widespread screening recommendations according to the US Preventive Task Force. In contrast, screening programs using photofluorography and upper endoscopy are prevalent in Asian countries like Japan and Korea. However, focused interventions could benefit certain subpopulations, such as Hispanic patients, which in the present study had significantly higher odds of late-stage presentation compared to their non-Hispanic Black, White, and Asian counterparts. There is evidence from previous studies that Hispanic patient populations have a significantly greater likelihood of late-stage presentation as well.^31^ However, the reasons for this association is not entirely clear. One explanation may be due to the disparity in insurance coverage among Hispanic patients in the US,^29^ leading to the lack of access to healthcare, with Hispanic patients being estimated to be the largest uninsured population. Therefore, research is needed to explore this further.

The present study, however, is not without limitations. SEER does not provide information on the timing of insurance coverage. Prior research demonstrated that cancer patients who were on Medicaid prior to their diagnosis had better survival than those enrolled at the time of diagnosis. Therefore, patients who were qualified for Medicaid only around their diagnoses might not have been accurately classified in this study. Second, the SEER database also limited the control of collected data and how variables were measured. For example, comorbidity, environmental exposures, molecular phenotypes, adjuvant treatments, family history, and lifestyle risk factors were not collected. Comorbid conditions could present concomitantly at the time of cancer diagnosis and have been found to affect survival rates. Therefore, these potential confounders could affect the associations we observed in this study, and thus, future studies should take into account these unmeasured factors. Lastly, the potential for misclassification of insurance status could not be avoided due to the way in which participants were inherently enrolled. Despite these limitations, the strength of our study lies in its large sample size and population-based design, with racially/ethnically diverse patient cohort in the SEER database. A large population-based study affords greater power to detect true differences compared to prior studies with small sample sizes.

In conclusion, our study showed that compared with privately insured GC patients, those without insurance or on Medicaid had worse GC-specific survival and tended to present at more advanced stage of GC. Furthermore, Hispanic patients had a later stage of disease at presentation than other racial/ethnic groups. Racial/ethnic minority patients had a similar survival rate than their non-Hispanic White counterparts. Together, these findings highlight the potential need for policy interventions addressing health insurance coverage, especially for Hispanic and other minority ethnic groups. In addition, the results of this study could be useful in developing public health strategies that target the development of appropriate screening for minority ethnic groups and other subpopulations at risk of more severe outcomes from GC.

## Data Availability

All data produced are available online at the Surveillance Epidemiology and End Results (SEER), 18 Registry.

https://seer.cancer.gov/registries/terms.html

## Notes

### Competing Interest Statement

The authors have declared no competing interest.

### Funding Statement

This work was supported in part by Susan G. Komen (TREND21675016) and the National Institute on Aging under grant award T32AG000243. Its contents are solely the responsibility of the author(s) and do not necessarily represent the official views of the National Institutes of Health/National Institute on Aging.

### Author Declarations

The study used publicly available data from the Surveillance Epidemiology and End Results (SEER), 18 Registry.

